# Association between Cardiovascular Burden and Requirement of Intensive Care among Patients with Mild COVID-19

**DOI:** 10.1101/2020.05.25.20111757

**Authors:** Shi Tai, Jianjun Tang, Bilian Yu, Liang Tang, Yang Wang, Huilin Zhang, Weihong Zhu, Kui Xiao, Chuan Wen, Chongqin Tan, Zhongbiao Jiang, Chuanhao Jiang, Li Zhu, Li Jiang, Qiming Liu, Xinqun Hu, Zhenfei Fang, Xuping Li, Jiaxing Sun, Zhaowei Zhu, Hui Yang, Tao Tu, Yichao Xiao, Mingxian Chen, Yuhu He, Xiangping Chai, Junmei Xu, Shenghua Zhou

## Abstract

**Background:** Information regarding the impact of cardiovascular disease (CVD) on disease progression among patients with mild coronavirus disease 2019 (COVID-19) is limited.

**Methods:** This study evaluated the association of underlying CVD with disease progression in patients with mild COVID-19. The primary outcome was the need to be transferred to intensive care due to disease progression. The patients were divided with and without CVD as well as stable and intensive care groups.

**Results:** Of 332 patients with mild COVID-19, median age was 51 years (IQR, 40-59 years), and 200 (61.2%) were female. Of 48 (14.5%) patients with CVD, 23 (47.9%) progressed to severe disease status and required intensive care. Compared with patients without CVD, patients with CVD were older, and more likely to have fatigue, chest tightness, and myalgia. The rate of requiring intensive care was significantly higher among patients with CVD than in patients without CVD (47.92% vs. 12.4%; P<0.001). In subgroup analysis, rate of requiring intensive care was also higher among patients with either hypertension or coronary heart disease than in patients without hypertension or coronary heart disease. The multivariable regression model showed CVD served as an independent risk factor for intensive care (Odd ratio [OR], 2.652 [95% CI, 1.019-6.899]) after adjustment for various cofounders.

**Conclusions:** Patients with mild COVID-19 complicating CVD in are susceptible to develop severe disease status and requirement for intensive care.

**Key Points:** *Question:* What is the impact of coexisting cardiovascular diseases (CVD) on disease progression in patients with mild COVID-19?

*Findings:* Although most patients with mild COVID-19 were discharged alive from hospital, approximately 47.9% patients with coexisting CVD developed severe disease status and required intensive care. CVD is an independent risk factor of intensive care among patients with mild COVID-19.

*Meaning:* Coexisting CVD is associated with unfavorable outcomes among patients with mild COVID-19. Special monitoring is required for these patients to improve their outcome.

## Introduction

The outbreak of coronavirus disease 2019 (COVID-19), which caused by severe acute respiratory syndrome coronavirus 2 (SARS-CoV-2) virus,^1^ rapidly progressed to a pandemic, and was declared a public health concern.^2^ Genetic sequencing of the virus suggests that SARS-CoV-2 is a betacoronavirus closely linked to the SARS virus. While most people with COVID-19 would develop mild or uncomplicated illness, approximately 14% people develop severe disease and finally require to be hospitalized to receive oxygen support, unfortunately, 5% patients even need to be treated in intensive care unit.^3, 4^ Importantly, a large proportion of affected patients have been reported to have underlying cardiovascular diseases (CVD),^5, 6^ and can present additional challenge in the battle against outbreaks of novel virus infections. However, information on factors affecting disease progression among patients with mild COVID-19 is limited.

Coronaviruses are known to affect the cardiovascular system,^7^ and the prevalence of underlying CVD is common and steadily increasing in China during recent decades, which may serve as a risk factor to the susceptibility and severity of infectious diseases.^8^ Although most mild patients with COVID-19 are thought to have a favorable prognosis, those with chronic underlying conditions may suffer from worse outcomes.^9^ Nevertheless, there is sparse data on clinical presentation of COVID-19 in specific populations, such as mild illness and coexisting CVD. In present study, we report the clinical characteristics and factors associated with developing severe disease after admission in patients with mild COVID-19.

## Methods

### Study Participants, criteria for patient admission, discharge, and referral

Consecutive patients admitted to temporary hospital with laboratory-confirmed COVID-19 from February 5, 2020, to March 10, 2020 were included in this retrospective cohort study. The criteria for patient admission, discharge, and referral were established according to World Health Organization interim guidance4 and Guidance for Corona Virus Disease 2019: Prevention, Control, Diagnosis and Management (Tentative Sixth Edition).^10^

Patients who met all the following conditions were discharged alive from hospital: (1) normal temperature for more than 3 consecutive days; (2) significantly improved respiratory symptoms; (3) resting SpO_2_ >93%; (4) radiology examination suggesting a significantly reduced pulmonary inflammation; (5) negative results of two consecutive SARS-CoV-2 testing by realtime reverse transcription–polymerase chain (RT-PCR) within 24-hour interval.

Patients with one of the following conditions were referred to the designated hospital for intensive care: (1) respiratory rate ≥ 30 breaths per minute; (2) resting SpO_2_ ≤ 93%; (3) temperature ≥ 38.5°C for more than 2 consecutive days even with proper treatment; (4) severe dysfunction of the heart, liver, lung, kidney, or brain; (5) other emergent conditions.

This study was approved by the National Health Commission of China and the Institutional Review Board at Renmin Hospital of Wuhan University (Wuhan, China).Written informed consent was waived by the ethics commission of the designated hospital for patients with emerging infectious diseases.

### Data Collection

The demographic characteristics (age and sex), clinical data (symptoms, comorbidities, laboratory findings, and outcomes) for participants during hospitalization were collected from electronic medical records by 2 investigators. The radiologic assessments included chest radiography or computed tomography. All data were independently reviewed and entered into the computer database by 2 analysts. Patients were categorized according to the presence or absence of CVD. CVD (cardiovascular diseases) include: hypertension, coronary artery disease, cerebrovascular disease (stroke), peripheral vascular disease, heart failure, rheumatic heart disease, congenital heart disease, and cardiomyopathies.

### Treatment

All patients with mild COVID-19 were treated according to the Guidance for Corona Virus Disease 2019: Prevention, Control, Diagnosis and Management (Tentative Sixth Edition). Antiviral therapy with umifenovir (0.2 g, three times daily for 5 days) or oseltamivir (75 mg, twice daily for 5 days) was administered. Traditional Chinese medicine with the Pneumonia Number 1 Decoction or Pneumonia Number 2 Decoction was also used for 3 days as a cycle until the symptoms were relieved. Statins, beta-blockers, and aspirin were administered for patients with coronary heart disease; Nifedipine was administered for patients with hypertension.

### Outcome

The clinical outcomes (ie, discharges, referral to the designated hospital for intensive care, and length of stay) were monitored up to March 25, 2020, the final date of follow-up. The end point was incidence of COVID-19 developing severe diseases status and requirement for intensive care. Successful treatment toward hospital discharge comprised relieved clinical symptoms, normal body temperature, significant resolution of inflammation as shown by chest radiography, and at least 2 consecutive negative results shown by RT-PCR chain reaction assay for SARS-CoV-2.

### Statistical Analysis

Descriptive statistics were obtained for all study variables. All categorical variables were compared for the study outcome by using the Fisher’s exact test or χ^2^ test, and continuous variables were compared using the t test or the Mann-Whitney U test, as appropriate. Continuous data are expressed as mean (SD) or median (interquartile range [IQR]) values. Categorical data are expressed as proportions. Survival curves were plotted using the Kaplan-Meier method and compared between patients with vs. without CVD using the log-rank test. Multivariate logistic regression models were used to determine the independent risk factors for referral during hospitalization. Data were analyzed using Stata15. Statistical charts were generated using Excel 2016 (Microsoft) or Stata15. For all the statistical analyses, P < 0.05 was considered significant.

## Results

### Demographics and Characteristics

Figure 1 shows the flowchart for patient recruitment. Briefly, of all 394 patients in the medical record system, who were screened initially from February 5, 2020, to March 10, 2020, 43 patients without available medical information and duplicated records, and 19 patients who transferred to designated hospital because of non-COVID-19 clinical factors were excluded. Finally, the study population consisted of 332 patients hospitalized with confirmed COVID-19: 58 patients (17.5%) required referral because requirement for intensive care (intensive group) and 274 patients (82.5%) discharged (stable group). The median age was 51 years (median [IQR] age, IQR, 40-59 years), and 200 (61.2%) were female. Among these patients, fever (177 patients [63.2%]) was the most common symptom. Cough, fatigue, chest tightness, and myalgia were present in 108 patients (38.4%), 32 patients (11.5%), 39 patients (13.9%),, and 12 patients (4.3%), respectively. Diarrhea (11 patients [3.9%]), sore throat (4 patients [1.4%]), rhinorrhea (4 patients [1.4%]), headache (7 patients [2.5%]), and toothache (2 patients [0.6%]) were rare. Hypertension (37 patients [11.1%]) and diabetes (11 patients [3.3%]) were the most common coexisting conditions. Table 1 presented clinical profile of stable patients and intensive care patients. Results showed that prevalence of chest tightness, toothache, CVD, coronary heart disease, hypertension, twice or more positive SARS-CoV-2 testing was significantly higher, while positive radiology findings was significantly lower in intensive care group than in stable group.

**Figure 1:**
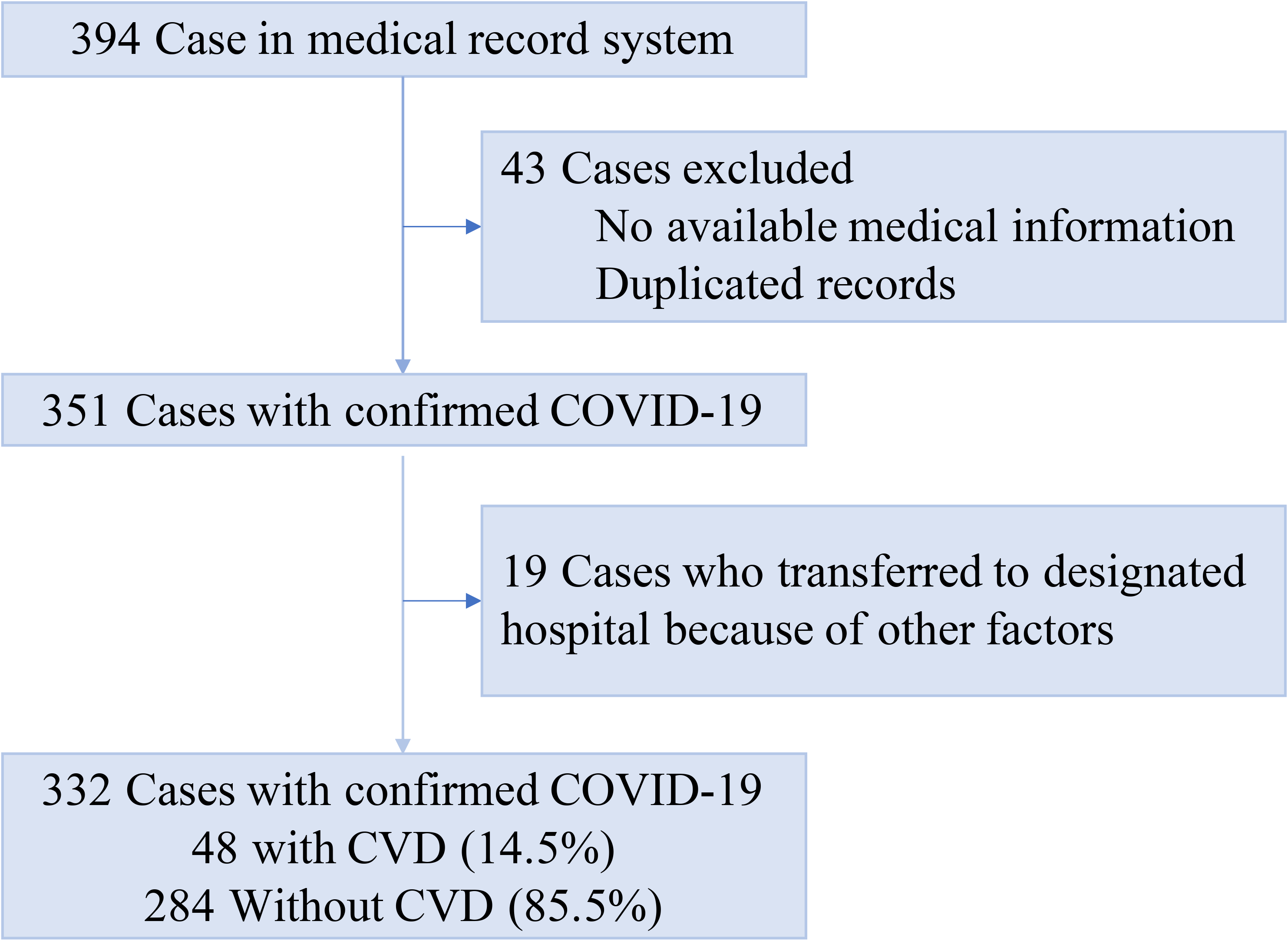
Flowchart of patient recruitment

**Table 1.** Clinic features of patients with mild COVID-19.

| Characteristic | All Patients<br>(n=332) | Stable group<br>(n=274, 82.5%) | Intensive care group<br>(n=58, 17.5%) | P Value |
| --- | --- | --- | --- | --- |
| **Sex** |  |  |  |  |
| Male, n (%) | 132 (39.8) | 105 (38.3) | 27 (46.6) | 0.245 |
| Female, n (%) | 200 (61.2) | 169 (61.7) | 31 (53.4) | 0.245 |
| **Age, median (IQR)** | 51 (40-59) | 50.5 (40-59) | 52 (42-60) | 0.478 |
| **Symptoms** |  |  |  |  |
| Fever, n (%) | 177 (63.2) | 142 (63.4) | 35 (62.5) | 0.901 |
| Cough, n (%) | 108 (38.4) | 88 (39.1) | 20 (35.7) | 0.640 |
| Fatigue, n (%) | 39 (13.9) | 28 (12.5) | 11 (19.6) | 0.167 |
| Chest tightness, n (%) | 32 (11.5) | 15 (6.7) | 17 (29.3) | <0.001 |
| Diarrhea, n (%) | 11 (3.9) | 8 (3.6) | 3 (5.4) | 0.538 |
| Headache, n (%) | 7(2.5) | 6(2.7) | 1(1.8) | 0.702 |
| Sore throat, n (%) | 4(1.4) | 2(0.9) | 2(3.4) | 0.181 |
| Rhinorrhea, n (%) | 4(1.4) | 4(1.8) | 0(0.0) | 0.587 |
| Myalgia, n (%) | 12(4.3) | 9(4.0) | 3(5.4) | 0.658 |
| Toothache, n (%) | 2 (0.6) | 0 (0.0) | 2 (3.4) | 0.030 |
| **Comorbidity** |  |  |  |  |
| CVD, n (%) | 48(14.5) | 25(7.5) | 23(39.7) | <0.001 |
| Coronary heart disease, n (%) | 11(3.3) | 1(0.4) | 10(17.2) | <0.001 |
| Hypertension, n (%) | 38(11.4) | 21 (7.7) | 17 (27.9) | <0.001 |
| Diabetes, n (%) | 11 (3.3) | 7 (2.6) | 4 (6.9) | 0.106 |
| Lung disease, n (%) | 7 (2.1) | 4 (1.5) | 3 (5.2) | 0.105 |
| **Radiologic abnormality, n (%)** | 201 (65.5) | 183 (70.1) | 18 (36.7) | <0.001 |
| **Positive SARS-CoV-2 testing by RT-PCR** |  |  |  |  |
| Twice or more, n (%) | 72 (23.0) | 48 (18.3) | 24 (48.0) | <0.001 |
| Once, n (%) | 241 (77.0) | 215 (81.7) | 26 (74) | <0.001 |
| **Laboratory findings** |  |  |  |  |
| Red-blood cell count*10<sup>12</sup>/L, median (IQR) | 4.39 (4.05-4.73) | 4.39(4.05-4.73) | 4.42(4.02-4.76) | 0.863 |
| White-cell count *10<sup>9</sup>/L, median (IQR) | 5.25 (4.49-6.37) | 5.19(4.47-6.33) | 5.62(4.88-6.58) | 0.058 |
| Neutrophil count *10<sup>9</sup>/L, median (IQR) | 3.21 (2.57-4.08) | 3.18(2.55-3.98) | 3.49(2.67-4.27) | 0.143 |
| Lymphocyte count, *10<sup>9</sup>/L, median (IQR) | 1.61 (1.26-1.91) | 1.58(1.26-1.86) | 1.80(1.26-2.17) | 0.057 |
| NLR, median (IQR) | 2.1(1.6-2.7) | 2.1 (1.6-2.7) | 2.1 (1.5-2.8) | 0.911 |
| Platelet count*10<sup>9</sup>/L, median (IQR) | 258 (216-310) | 267(218-324) | 223(194-269) | 0.003 |
| Hemoglobin, g/L, median (IQR) | 135 (125-145) | 135(126-144) | 135(122-145) | 0.781 |
| C-reactive protein |  |  |  |  |
| <10.00 mg/L | 261 (85.0) | 217 (84.4) | 44 (88.0) | 0.518 |
| ≥10.00 mg/L | 46 (15.0) | 40 (15.6) | 6 (12.0) | 0.518 |
| **Median length of symptoms onset to admission, days**<br>**(IQR)** | 12 (8-16) | 12 (8-15) | 13 (6-21) | 0.679 |
| **Median length of hospital stay, days**<br>**(IQR)** | 14 (10-19) | 14 (10-19) | 15 (11-21.5) | 0.322 |

Abbreviation: IQR, interquartile range; NLR, neutrophil to lymphocyte ratio.

Patients were also grouped as non-CVD and CVD patients. Compared with patients without CVD, patients with CVD were older (56 [45-62] years vs. 50 [39-59] years; P=0.007), and more likely to have fatigue (13 of 48 patients [28.3%] vs. 26 of 284 patients [11.1%]; P=0.002), chest tightness (18 of 48 patients [40.0%] vs. 14 of 284 patients [6.0%]; P <0.001), and myalgia (6 of 48 patients [13.0%] vs. 6 of 284 patients [2.6%]; P=0.001).Moreover, comorbidities, including diabetes (4 [8.3%] vs. 7 [2.5%]) and lung disease (4 [8.3%] vs. 3 [1.1%]) were present more often among patients with CVD (all P<0.05) (Table 2). In subgroup analysis, compared with patients without hypertension, patients with hypertension were older (56 [49-60] years vs. 50 [39-59] years; P=0.009), and more likely to have fatigue (10 of 38 patients [27.0%] vs. 29 of 294 patients [11.9%]; P=0.014), and chest tightness (14 of 38 patients [38.9%] vs. 18 of 294 patients [7.4%]; P <0.001). Moreover, comorbidities, including diabetes (4 [10.5%] vs. 7 [2.4%]; P=0.027) was present more often among patients with hypertension; Compared with patients without coronary heart disease, patients with coronary heart disease more likely to have fatigue (5 of 11 patients [45.5%] vs. 34 of 321 patients [12.6%]; P=0.002), and chest tightness (6 of 11 patients [54.5%] vs. 26 of 321 patients [9.7%]; P <0.001). Lung disease (2 [18.2%] vs. 5 [1.6%]; P=0.019) was present more often among patients with coronary heart disease (Supplemental Table 1).

**Table 2.** Clinic characteristic of patients with COVID-19 according to with or without CVD.

| Characteristic | All patients<br>(n=332) | Without CVD<br>(n=284, 85.5%) | With CVD<br>(n=48, 14.5%) | *P* value |
| --- | --- | --- | --- | --- |
| **Sex** |  |  |  |  |
| Male, n (%) | 132 (39.8) | 110 (38.7) | 22 (45.8) | 0.285 |
| Female, n (%) | 200 (60.2) | 174 (61.3) | 26 (54.2) | 0.285 |
| **Age, median (IQR)** | 51 (40-59) | 50 (39-59) | 56 (45-62) | 0.007 |
| **Symptoms** |  |  |  |  |
| Fever, n (%) | 177 (63.2) | 150 (63.3) | 27 (58.7) | 0.487 |
| Cough, n (%) | 108 (38.4) | 88 (37.6) | 20 (42.6) | 0.525 |
| Fatigue, n (%) | 39 (13.9) | 26 (11.1) | 13 (28.3) | 0.002 |
| Chest tightness, n (%) | 32(11.5) | 14(6.0) | 18(40) | <0.001 |
| Diarrhea, n (%) | 11(3.9) | 9(3.8) | 2(4.3) | 0.873 |
| Headache, n (%) | 7(2.5) | 5(2.1) | 2(4.3) | 0.323 |
| Sore throat, n (%) | 4(1.4) | 4(1.7) | 0(0.0) | 1.000 |
| Rhinorrhea, n (%) | 4(1.4) | 4(1.7) | 0(0.0) | 1.000 |
| Myalgia, n (%) | 12(4.3) | 6(2.6) | 6(13.0) | 0.001 |
| Toothache, n (%) | 2 (0.6) | 1 (0.4) | 1 (2.1) | 0.269 |
| **Comorbidity** |  |  |  |  |
| Diabetes, n (%) | 11 (3.3) | 7 (2.5) | 4 (8.3) | 0.036 |
| Lung disease , n ( % ) | 7 (2.1) | 3 (1.1) | 4 (8.3) | 0.010 |
| **Radiologic abnormality, n (%)** | 201 (65.5) | 182 (67.2) | 19 (52.8) | 0.088 |
| **Positive SARS-CoV-2 testing by RT-PCR** |  |  |  |  |
| Twice or more, n (%) | 72(23.0) | 60(22.0) | 12(35.1) | 0.147 |
| Once, n (%) | 241(77.0) | 216(78.0) | 25(64.9) | 0.147 |
| **Laboratory findings** |  |  |  |  |
| Red-blood cell count*10<sup>12</sup>/L, median (IQR) | 4.39 (4.05-4.73) | 4.37 (4.04-4.68) | 4.49 (4.15-4.90) | 0.159 |
| White-cell count *10<sup>9</sup>/L, median (IQR) | 5.25 (4.49-6.37) | 5.25 (4.49-6.37) | 5.00 (4.46-6.35) | 0.677 |
| Neutrophil count *10<sup>9</sup>/L, median (IQR) | 3.21 (2.57-4.08) | 3.23 (2.59-4.10) | 3.06 (2.53-3.91) | 0.468 |
| Lymphocyte count*10<sup>9</sup>/L, median (IQR) | 1.61 (1.26-1.91) | 1.58 (1.27-1.91) | 1.72 (1.23-1.92) | 0.745 |
| NLR, median (IQR) | 2.06 (1.60-2.67) | 2.07 (1.60-2.67) | 2.04 (1.56-2.54) | 0.609 |
| Platelet count*10<sup>9</sup>/L, median (IQR) | 258 (216-310) | 263 (217-314) | 243 (202-287) | 0.085 |
| Hemoglobin, g/L, median (IQR) | 135 (125-145) | 135 (125-145) | 137 (130-145) | 0.254 |
| C-reactive protein, |  |  |  |  |
| <10.00 mg/L | 261(85.0) | 230(85.8) | 31(79.5) | 0.300 |
| ≥10.00 mg/L | 46(15.0) | 38(14.2) | 8(20.5) | 0.300 |
| **Median length of symptoms onset to admission, days (IQR)** | 12(8-16) | 12(8-15) | 14(8-18) | 0.222 |
| **Median length of hospital stay, days (IQR)** | 14(10-19) | 14(10-19) | 15(8-20) | 0.837 |
| **Clinical outcomes at data cutoff** |  |  |  |  |
| Discharge, n (%) | 274(82.5) | 249(87.6) | 25(52.1) | <0.001 |
| Require intensive care, n (%) | 58(17.5) | 35(12.4) | 23(47.9) | <0.001 |

Abbreviation: IQR, interquartile range; NLR, neutrophil to lymphocyte ratio.

### Laboratory and Radiographic Findings

In terms of laboratory findings, there were no significantly statistic differences between patients with and without CVD, including white-cell count, neutrophil count, neutrophil to lymphocyte ratio (NLR), lymphocyte count, platelet count, and C-reactive protein. Results of SARS-CoV-2 testing and radiologic findings were similar between 2 groups. In subgroup analysis, patients with coronary heart disease vs. those without coronary heart disease had better radiologic results on admission.

### Clinical Outcome

Of 332 patients, 48 (14.5%) had underlying CVD including hypertension and coronary heart disease, and 23 (47.9%) developed severe diseases who required intensive care. The median time from symptom onset to admission was 12 days (IQR, 8-16 days) and median duration of hospital stay was 14 days (IQR, 10-19 days). Patients with CVD vs. those without CVD had longer durations from symptom onset to admission (mean, 14 [range, 8-18] days vs. 12 [range, 8-15] days) and duration of hospital stay (15 [IQR, 8-20] days vs. 14 [IQR, 10-19] days). As shown in Table 2 and the Kaplan-Meier curves in Figure 2, the requirement for intensive care was higher among patients with vs. without CVD (23 of 48 [47.9%] vs. 35 of 284 [12.4%]; P < 0.001). In subgroup analysis, the requirement for intensive care rate was higher among patients with vs without either hypertension or coronary heart disease (17 [44.7%] vs. 13 [13.9%]; 10 [90.9%] vs. 48 [15.0%]; all P< 0.001) as shown in Supplemental Table 1 and Kaplan-Meier curves in Supplemental Figure 1-2. AS shown in Supplemental Table 2, chest tightness, CVD, platelet count, radiologic abnormality, more than once positive SARS-CoV-2 testing by RT-PCR were risk factors of intensive care in this patient cohort. To evaluate whether CVD is associated with requirement for intensive care in patients with mild COVID-19, we adjusted for demographic characteristics, comorbidities, and clinical profiles. Model 1 adjusted for age and sex; Model 2 adjusted for age, sex, chest tightness, diabetes mellitus and lung diseases; Model 3 adjusted for age, sex, chest tightness, diabetes mellitus and lung diseases, platelet count, radiologic abnormality, and more than once positive SARS-CoV-2 testing by RT-PCR. As shown in Table 3, CVD remains as an independent risk factor for requirement for intensive care in patients with mild COVID-19 after adjustment for cofounders by Model 1, 2 and 3.

**Figure 2:**
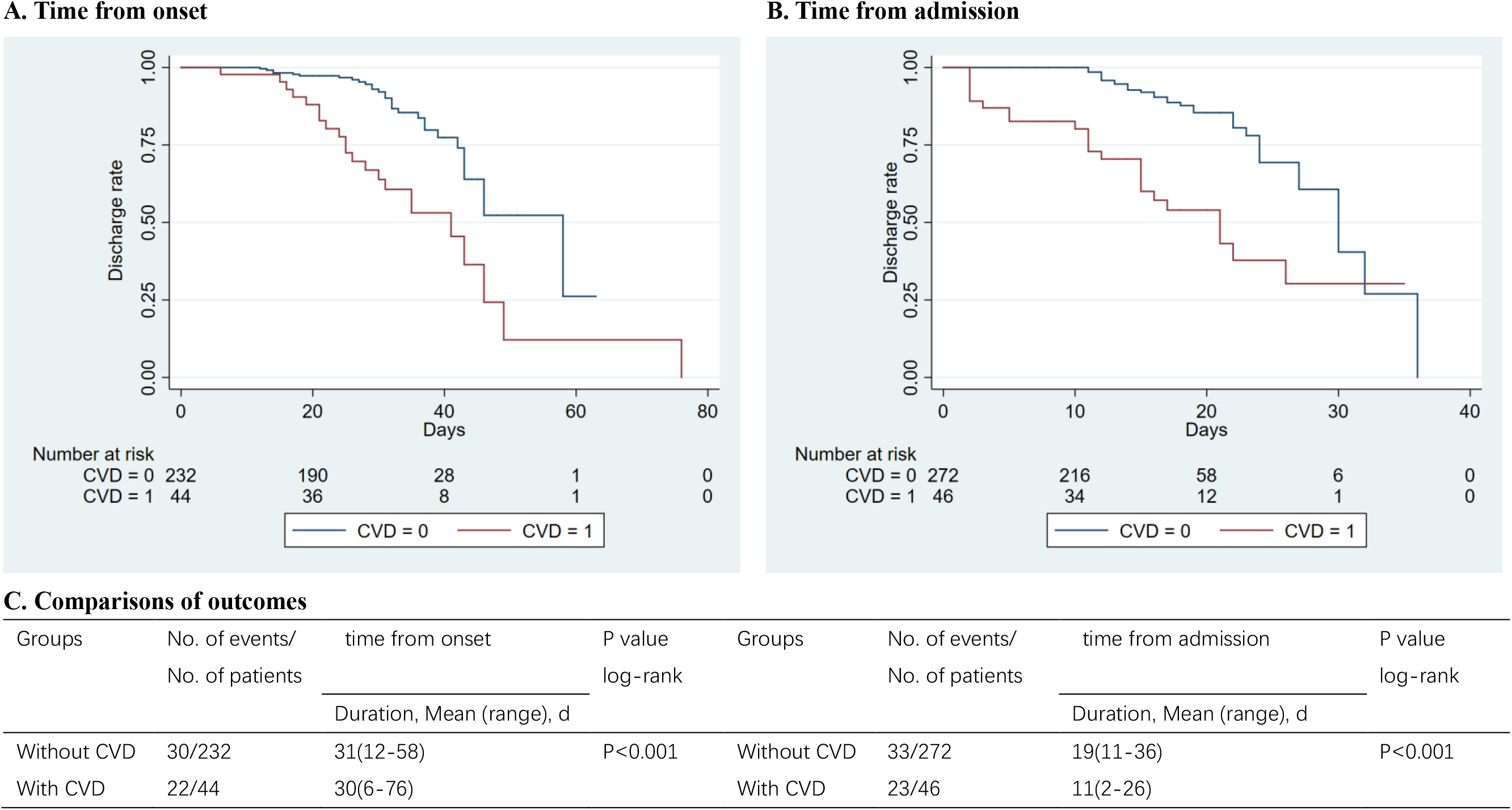
Discharge rate during hospitalization between patients with vs without CVD A-B, Kaplan-Meier survival curves for intensive care during the time from symptom onset (A) and admission (B). C, Patients with CVD had a higher rate of intensive care in log-rank test, both from symptom onset and from admission.

**Table 3.** Multivariable analyses of the risk factors for intensive care in patients with mild COVID-19.

|  | Unadjusted |  |  | Model 1 |  |  | Model 2 |  |  | Model 3 |  |  |
| --- | --- | --- | --- | --- | --- | --- | --- | --- | --- | --- | --- | --- |
|  | OR | 95% CI | *P* | OR | 95% CI | *P* | OR | 95% CI | *P* | OR | 95% CI | *P* |
| With CVD | 6.545 | 3.357-12.762 | 0.001 | 6.430 | 3.210-12.878 | *P*<0.001 | 3.941 | 1.798-8.636 | 0.001 | 2.652 | 1.019-6.899 | 0.046 |
| Age | 1.006 | 0.983-1.030 | 0.604 | 0.994 | 0.969-1.020 | 0.647 | 0.993 | 0.966-1.020 | 0.601 | 0.991 | 0.961-1.021 | 0.545 |
| Sex | 1.402 | 0.792-2.480 | 0.246 | 1.284 | 0.699-2.359 | 0.007 | 1.384 | 0.720-2.663 | 0.330 | 0.960 | 0.447-2.062 | 0.916 |
| Chest tightness | 6.044 | 2.788-13.107 | 0.001 |  |  |  | 3.451 | 1.422-8.376 | 0.006 | 3.340 | 1.070-10.429 | 0.038 |
| Diabetes mellitus | 2.825 | 0.799-9.987 | 0.107 |  |  |  | 2.473 | 0.606-10.087 | 0.207 | 2.784 | 0.604-12.826 | 0.189 |
| Lung diseases | 3.682 | 0.801-16.915 | 0.094 |  |  |  | 3.604 | 0.557-23.327 | 0.178 | 3.806 | 0.480-30.183 | 0.206 |
| Platelet | 0.991 | 0.986-0.996 | <0.001 |  |  |  |  |  |  | 0.233 | 0.075-0.726 | 0.012 |
| Radiologic abnormality | 0.238 | 0.125-0.451 | 0.001 |  |  |  |  |  |  | 0.218 | 0.101-0.468 | 0.001 |
| Positive SARS-CoV-2 testing by RT-PCR≥2 | 4.135 | 2.187-7.817 | 0.001 |  |  |  |  |  |  | 3.754 | 1.719-8.197 | 0.001 |

Sample size, n=332. Data are expressed as OR±95% CIs (reported in parentheses) as assessed by univariate (unadjusted) or multivariate logistic regression analyses.
OR: odds ratio; CI: confidence interval; CVD: cardiovascular diseases.
In these logistic regression models, CVD was a categorical variable.
Other covariates included in multivariate logistic regression models were model 1: adjustment for age and sex; model 2: adjustment for variables including age, sex, chest tightness, diabetes mellitus and lung diseases; model 3: adjustment for variables including age, sex, chest tightness, diabetes mellitus, lung diseases, platelet, radiologic abnormality, and positive SARS-CoV-2 testing by RT-PCR, platelet.

## Discussion

Since the incidence of cardiovascular disease is steadily increasing in China for decades.^11, 12^ Many hospitalized patients might naturally have comorbid CVD, recent observations that patients with preexisting CVD are susceptible to the most adverse complications among patients with severe COVID-19 diseases.^6, 13-15^ However, the impact of CVD on outcome of mild COVID-19 patients is limited. The present study demonstrates that comorbid CVD is an independent risk factor for the development of severe COVID-19 and requirement of intensive care among patients with mild COVID-19.

In present study, among hospitalized patients with mild COVID-19, fever, cough, and fatigue were the most common symptoms, which is consistent with previous reports.^14, 16^ Notably, in comparison with patients without CVD, patients with CVD showed significantly higher rates of chest tightness, fatigue, and myalgia. These finding suggests that patients with comorbidities had more severe symptoms, which clearly overlapped with the clinical symptoms of cardiovascular diseases. In the process of clinical practice, cautious screening is required to determine whether the symptoms are caused by COVID-19 or progression of underlying diseases. Particularly, chest tightness and fatigue might be associated with CVD including heart failure. Therefore, SARS-CoV-2 may induce and complicate the typical symptoms and manifestations of acute myocardial infarction and chronic heart failure.

Nationwide retrospective study showed that CVDs were the most common coexisting conditions in patients hospitalized with COVID-19.^15^ A meta-analysis of six studies including 1527 patients with COVID-19 examined the prevalence of CVD and reported the prevalence of hypertension, cardiac and cerebrovascular disease, and diabetes to be 17.1%, 16.4%, and 9.7%, respectively.16 In present study, 13.31% patients with mild cases had CVD. Compared to discharged patients, patients with progressive disease progression and referred to a designated hospital for intensive care were more likely to have coexisted CVD. Multivariate regression analysis suggested that CVD was an independent risk factor for patients developing severe disease after adjustment of various cofounders (Table 3). In subgroup analysis, although mild COVID-19 patients with CVD had less positive radiologic findings than patients without CVD on admission, they still faced higher risk of unfavorable disease progression as compared to patients without CVD. Regarding the possible mechanisms, prevalent CVD, as an inflammation disease, may serve as a marker of accelerated immunologic dysregulation and thus, aggravate the immunological imbalance induced by COVID-19. On the other hand, COVID-19 itself, might aggravate the myocardial injury, recent studies from Wuhan demonstrated severe COVID patients with the most ominous outcome had significantly higher TnI or TnT levels as compared to severe COVID-19 patients undergoing recovery^17,18^. It is known that SARS-CoV-2 can elicit the intense release of multiple cytokines and chemokines that can not only lead to vascular inflammation and plaque instability, but also to myocardial inflammation^6, 12^, which might be linked with the enhanced myocardial injury during the disease process. The interaction between preexisted CVD and the harm of SARS-CoV-2 on myocardial tissue might thus imply some of the mechanisms responsible for the worse outcome among mild COVID-19 patients hospitalized.

Among patients with mild cases, hypertension was the most common comorbidity, followed by coronary heart disease. An early analysis of underlying clinical features in 41 COVID-19 fatal cases revealed the highest prevalence of hypertension (60.9%).^6^ Higher expression of ACE2 in patients with hypertension has been postulated to enhance susceptibility to SARS-CoV2^19^, although the data are conflicting and without clear suggestion implication for treatment.^20^ Additional study is needed to understand the potential mechanistic relationships between hypertension and COVID-19 outcomes.

## Limitations

Our study has several limitations. First, restricted by the limited medical resources, laboratory biochemical testing was insufficient; second, regarding the time of disease onset in patients, the uncertainty of the exact dates (recall bias) might have inevitably affected our assessment; lastly, data from the earliest hospitalized patients could not be fully accessed.

## Clinical implication

Mild COVID patients with coexisting CVD should receive more intensive monitoring to prevent the transition severe disease progression.

## Conclusions

Although most mild COVID-19 patients could be discharged, approximately half of the mild COVID-19 patients with comorbid CVD may progress to severe disease status and require intensive care, special monitoring on these patient populations are essential to improve the outcome of these patients.

## Data Availability

The data used to support the findings of this study are included within the supplementary information file.

## Author Contributions

Drs S. Zhou and J. Xu had full access to all of the data in the study and take responsibility for the integrity of the data and the accuracy of the data analysis. Concept and design: S. Tai, J. Tang, B. Yu, L. Tang, Y. Wang, X. Chai, S. Zhou. Acquisition, analysis, or interpretation of data: S. Tai, J. Tang, B. Yu, L. Tang, S. Zhou. Drafting of the manuscript: S. Tai, J. Sun, L. Tang, S. Zhou. Critical revision of the manuscript for important intellectual content: Y. Wang, H. Zhang, K. Xiao, C. Wen, C. Tan, Z. Jiang, C. Jiang. Statistical analysis: S. Tai, J. Sun, L. Zhu, L. Jiang. Obtained funding: S. Tai, S. Zhou. Administrative, technical, or material support: Q. Liu, Z. Fang, X. Hu, X. Li, Z. Zhu, H. Yang, T. Tao, Y. Xiao, M. Chen, Y. He. Supervision: X. Chai, J. Xu, S. Zhou.

---

**Supplemental Figure 1:** A-B, Kaplan-Meier survival curves for intensive care during the time from symptom onset (A) and admission (B). C, Patients with hypertension had a higher rate of intensive care in log-rank test, both from symptom onset and from admission.

**Supplemental Figure 2:** A-B, Kaplan-Meier survival curves for intensive care during the time from symptom onset (A) and admission (B). C, Patients with coronary heart disease had a higher rate of intensive care in log-rank test, both from symptom onset and from admission.

## References

1. Zhu N, Zhang D, Wang W, et al. A Novel Coronavirus from Patients with Pneumonia in China, 2019. N Engl J Med. 2020;382:727–733.

2. WHO Director-General’s opening remarks at the media briefing on COVID-19 - 11 March 2020: World Health Organization.

3. Novel Coronavirus Pneumonia Emergency Response Epidemiology T. [The epidemiological characteristics of an outbreak of 2019 novel coronavirus diseases (COVID-19) in China]. Zhonghua Liu Xing Bing Xue Za Zhi. 2020;41:145–151.

4. Clinical management of severe acute respiratory infection (SARI) when COVID-19 disease is suspected: Interim guidance. World Health Organization 2020.

5. Guan WJ, Ni ZY, Hu Y, et al. Clinical Characteristics of Coronavirus Disease 2019 in China. N Engl J Med. 2020.

6. Huang C, Wang Y, Li X, et al. Clinical features of patients infected with 2019 novel coronavirus in Wuhan, China. Lancet. 2020;395:497–506.

7. Cowan LT, Lutsey PL, Pankow JS, Matsushita K, Ishigami J, Lakshminarayan K. Inpatient and Outpatient Infection as a Trigger of Cardiovascular Disease: The ARIC Study. J Am Heart Assoc. 2018;7:e009683.

8. Dhainaut JF, Claessens YE, Janes J, Nelson DR. Underlying disorders and their impact on the host response to infection. Clin Infect Dis. 2005;41 Suppl 7:S481–489.

9. Zhou F, Yu T, Du R, et al. Clinical course and risk factors for mortality of adult inpatients with COVID-19 in Wuhan, China: a retrospective cohort study. Lancet. 2020.

10. Guidance for Corona Virus Disease 2019: Prevention, Control, Diagnosis and Management: People’s Medical Publishing House; 2020.

11. Bonow RO, Fonarow GC, O’Gara PT, Yancy CW. Association of Coronavirus Disease 2019 (COVID-19) With Myocardial Injury and Mortality. JAMA Cardiol. 2020.

12. Yang C, Jin Z. An Acute Respiratory Infection Runs Into the Most Common Noncommunicable Epidemic-COVID-19 and Cardiovascular Diseases. JAMA Cardiol. 2020.

13. Chen N, Zhou M, Dong X, et al. Epidemiological and clinical characteristics of 99 cases of 2019 novel coronavirus pneumonia in Wuhan, China: a descriptive study. Lancet. 2020;395:507–513.

14. Wang D, Hu B, Hu C, et al. Clinical Characteristics of 138 Hospitalized Patients With 2019 Novel Coronavirus-Infected Pneumonia in Wuhan, China. JAMA. 2020.

15. Wu Z, McGoogan JM. Characteristics of and Important Lessons From the Coronavirus Disease 2019 (COVID-19) Outbreak in China: Summary of a Report of 72314 Cases From the Chinese Center for Disease Control and Prevention. JAMA. 2020.

16. Li B, Yang J, Zhao F, et al. Prevalence and impact of cardiovascular metabolic diseases on COVID-19 in China. Clin Res Cardiol. 2020.

17. Shi S, Qin M, Shen B, et al. Association of Cardiac Injury With Mortality in Hospitalized Patients With COVID-19 in Wuhan, China. JAMA Cardiol. 2020.

18. Guo T, Fan Y, Chen M, et al. Cardiovascular Implications of Fatal Outcomes of Patients With Coronavirus Disease 2019 (COVID-19). JAMA Cardiol. 2020.

19. Danser AHJ, Epstein M, Batlle D. Renin-Angiotensin System Blockers and the COVID-19 Pandemic: At Present There Is No Evidence to Abandon Renin-Angiotensin System Blockers. Hypertension. 2020:HYPERTENSIONAHA12015082.

20. Driggin E, Madhavan MV, Bikdeli B, et al. Cardiovascular Considerations for Patients, Health Care Workers, and Health Systems During the Coronavirus Disease 2019 (COVID-19) Pandemic. J Am Coll Cardiol. 2020.

